# ASSOCIATION BETWEEN QUADRICEPS STRENGTH AND KNEE FLEXION DURING DROP LANDING IN HEALTHY ADOLESCENT ATHLETES

**DOI:** 10.64898/2026.05.28.26353494

**Authors:** Brenden Lyons, Jackson Hopfauf, Colin W. Bond, Benjamin C. Noonan

**Affiliations:** Sanford Health, Fargo, North Dakota, USA

**Keywords:** ACL, Biomechanics, Drop Jump, Injury Risk, Stiffness

## Abstract

**Background:** Quadriceps strength and landing mechanics are two modifiable factors associated with anterior cruciate ligament (ACL) injury risk. Collecting detailed biomechanical data is an arduous task. Identifying a relationship using more easily measured variables, such as quadriceps strength, would offer value for athlete counseling and injury prevention programs. Although quadriceps weakness has been associated with altered landing strategies in ACL-reconstructed (ACLR) individuals, this relationship is less clear in healthy athletes.

**Purpose:** To investigate the association between isokinetic quadriceps strength and peak knee flexion angle during a vertical drop jump in healthy adolescent athletes.

**Study Design:** Secondary analysis of previously collected data.

**Methods:** Healthy adolescent athletes had their dominant leg quadriceps strength measured using an isokinetic dynamometer at 60°/s from 0–90° of knee flexion. Landing mechanics were assessed during a vertical drop jump using three-dimensional motion capture synchronized with force plates. Pearson correlation was used to evaluate the association between quadriceps strength and peak knee flexion angle during landing, with statistical significance defined as p < .05.

**Results:** There was a weak negative correlation between quadriceps strength and peak knee flexion angle (p = .017, R = -.22 [-.04, -.38]), suggesting that stronger athletes achieved greater knee flexion angles.

**Discussion:** Greater quadriceps strength was associated with increased peak knee flexion angles during landing; however, the weak correlation suggests that strength explains only a small portion of the variability in landing mechanics. These findings deviate slightly from prior literature in healthy populations but are consistent with studies demonstrating that greater quadriceps strength is associated with achieving greater peak knee flexion in ACLR patients. Accordingly, quadriceps strengthening should remain a key component of multifactorial ACL injury prevention programs.

## INTRODUCTION

There are many modifiable factors contributing to anterior cruciate ligament (ACL) injury risk with two important aspects being muscular strength and force attenuation strategy during landing tasks. It is widely thought these two characteristics are related to quadriceps strength that may influence sagittal plane knee movement while landing. The vertical drop jump test has been frequently used to assess sagittal plane mechanics to screen an individual’s risk for injury. Specifically, it has been shown that stiffer landings, those with less knee flexion and higher vertical ground reaction force (vGRF), are associated with an increased risk of ACL injury in young female athletes (1). Conversely, less stiff landings, those with more knee flexion, are associated with improved shock absorption and reduced tibial shear forces, thus potentially decreasing ACL injury risk (2). It has been postulated that sufficient eccentric strength of the quadriceps is important for controlling eccentric knee flexion during these landings.

There is a common assumption that athletes with weaker quadriceps have stiffer landings due to a conscious or subconscious inability or unwillingness to eccentrically control knee flexion; though, the relationship between these two variables may be more nuanced. Individuals recovering from ACL reconstruction (ACLR) with weaker thigh musculature are more likely to have less sagittal plane knee angle excursion during landing, which is mechanically associated with a stiffer landing (3). Mechanically, this landing is characterized by less knee flexion, thus shifting the role of energy dissipation from eccentrically controlled knee flexion to passive structures at the knee joint, such as tendons and ligaments or the bony alignment of the tibia and femur themselves. Yet, uninjured elite athletes also exhibit similar stiff landing patterns, even when it can be assumed they have high relative quadriceps strength (4). These stiffer landing patterns are associated with reduced ground contact time and increased impulse, resulting in greater acceleration during change of direction tasks (5). Therefore, factors other than quadriceps strength may have a more vital role in shaping landing kinematics related to force attenuation within a healthy athlete population. Understanding the relationship between quadriceps strength and landing mechanics is important because it can lead to more sensitive ACL injury risk screens, potentially leading to improved ACL injury prevention initiatives such as personalized training programs.

The purpose of this study is to investigate whether isokinetic quadriceps strength on the dominant leg, quantified as peak torque at 60°/s through 0 to 90° of knee flexion, is associated with greater peak knee flexion angles on the dominant leg during the landing phase of a drop jump in healthy adolescent athletes. We hypothesize that greater quadriceps strength will be moderately correlated with increased knee flexion during landing.

## METHODS

### Study Design and Participants

The present investigation is an observational, cross-sectional, secondary analysis of data previously collected as part of the 2024 Baseline ACL Injury Risk Screening and Normative Data study (ClinicalTrials.gov ID: NCT06635668). One hundred and thirty-four healthy adolescent athletes were originally enrolled in the ACL BUS 2024 cohort. Eligible athletes were free of injuries that limited full participation and reported no suspicion of pregnancy. All participants demonstrated at least a recreational activity level, with many engaged in competitive athletics. The study was approved by the Sanford Health Institutional Review Board (Study ID: 2976). Written informed consent or assent was obtained from all athletes, and parental consent was secured for minors.

### Data Collection Procedures

Testing took place at high schools located in the Upper Midwest. Upon enrollment, each athlete’s height, weight, and dominant leg were documented. Dominance was determined by asking participants which leg they would use to kick a soccer ball. Before beginning the injury risk screening, athletes completed a warm-up as part of sport practice or strength training. The screening protocol included the assessment of quadriceps strength with an isokinetic dynamometer and the evaluation of movement quality during a drop jump.

Muscle strength testing was performed using an isokinetic dynamometer (Humac-Norm, CSMi, Stoughton, MA). Each participant was properly positioned and secured in the apparatus, with administrators providing standardized instructions. Athletes were seated fully back in the chair, maintaining lumbar spine contact with the backrest, which was reclined to approximately 85° from horizontal. The backrest was adjusted such that, at 90° of knee flexion, a ∼0.5-inch space remained between the posterior shank and the seat edge. The chair and dynamometer arm were aligned so that the knee joint axis matched the dynamometer’s rotational axis. The lever arm pad was placed against the distal shank, just above the malleoli, and secured with hook-and-loop straps. A thigh strap was positioned across the distal posterior thigh of the testing leg, while the opposite limb was left unsupported. Participants were instructed to remain in contact with the backrest and hold the device’s handles throughout testing. The motion arc was standardized from full extension (0°) to 90° flexion, with the anatomical reference set at 90°. Each athlete performed a preliminary movement to ensure comfort and verify the absence of mechanical obstruction.

Testing consisted of two sets of reciprocal concentric knee flexion and extension at 60°/s for both legs. The first set, comprising five repetitions, served as a warm-up and familiarization trial, executed at progressively increasing submaximal effort. After a 20-second rest, a second set of five maximal-effort repetitions was performed through the complete range of motion. Test administrators provided consistent instructions and real-time verbal encouragement. Limb testing orders were alternated across participants to mitigate order effects and reduce equipment adjustments. Repetitions were required to be smooth, complete, and demonstrate sustained maximal effort; if criteria were not met, the set was repeated after a brief rest. Peak instantaneous concentric quadriceps torque on the dominant limb scaled to body mass was calculated.

For the drop jump, participants began each attempt standing upright on a 12-inch (0.30 m) plyometric box, placed six inches (0.15 m) behind the leading edge of the force plates. At the cue, they stepped forward with both feet simultaneously, landed bilaterally on the plates, and immediately executed a maximal vertical jump. Athletes were instructed to avoid stepping off with a single leg to reduce asymmetrical landing strategies, and to jump straight up as high and quickly as they could. No additional coaching was provided. One practice trial was completed, with additional practice allowed, if necessary, followed by two successful test trials. A trial was deemed invalid if the athlete led with a single leg, failed to land entirely on the plates, contacted both plates with one foot, or stepped off the plates before completing the vertical jump.

Motion data were captured with a Qualisys Miqus Video 3D motion capture system (Qualisys AB, Gothenburg, Sweden) at 110 Hz, synchronized with ground reaction force data from dual side-by-side force plates (Model 4060-07, Bertec Corporation, Columbus, OH, USA) sampled at 880 Hz. Three-dimensional pose reconstruction was performed using Theia3D (Version 2023.1.0.3160 p18, Theia Markerless Inc., Kingston, ON, Canada) with its integrated inverse kinematics model. Kinematic and kinetic processing was conducted in Visual3D (Version 2024.06.1, HAS-Motion Inc., Kingston, ON, Canada). Peak knee flexion angle for the dominant leg was extracted during the landing phase, defined from initial contact (vertical GRF >10 N) until toe-off (vertical GRF <10 N), and values were averaged.

### Statistical Analysis

The primary variables were peak knee flexion angle during the drop jump landing phase on the dominant leg and peak isokinetic concentric torque, or quadriceps strength, on the dominant leg. Quadriceps strength values were normalized to body mass to account for inter-subject variability. Pearson correlation was employed to examine the linear relation between quadriceps strength and peak knee flexion angle. A priori sample size calculation indicated that a minimum of 85 athletes would be required to detect a moderate correlation (r = 0.30) with 80% power at a two-sided significance level of 0.05.

## RESULTS

The Pearson correlation analysis revealed a weak negative correlation between quadriceps strength and peak knee flexion angle (p = .017, R = -.22 [-.04, -.38]), suggesting that stronger athletes achieved greater knee flexion angles.

**Figure 1.**
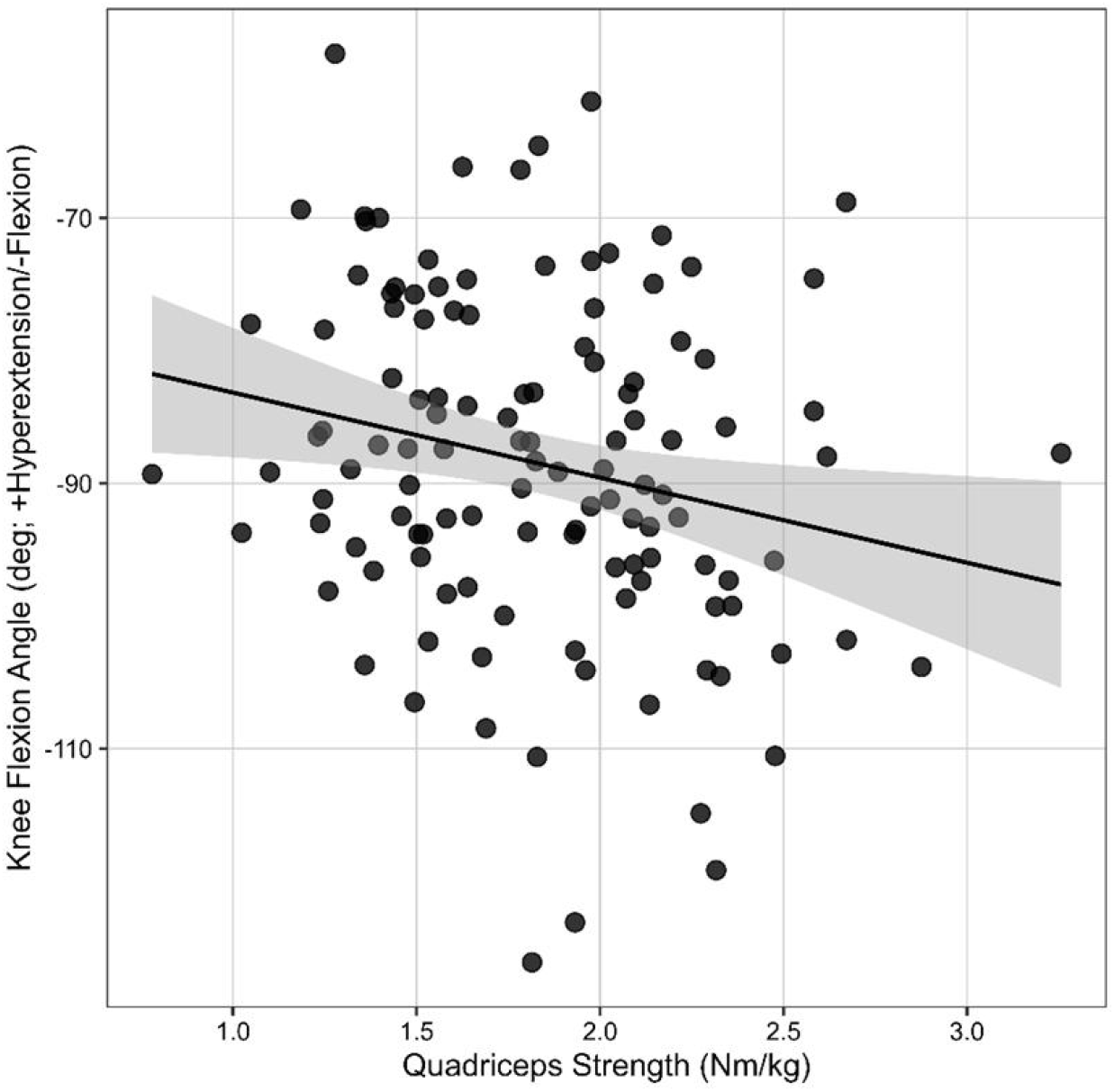
Relation between peak knee flexion angle and quadriceps strength. Black line: linear model; ribbon: 95% confidence interval for the linear model; black dots: individual subjects.

## DISCUSSION

The purpose of this investigation was to evaluate the association between isokinetic quadriceps strength and peak knee flexion angles in a vertical drop jump in healthy adolescent athletes. The results of this study support our hypothesis that athletes with stronger quadriceps will achieve a greater peak knee flexion angle. The key takeaway is that quadriceps strength relates to landing strategy in healthy adolescents, but its influence appears limited, suggesting that other factors play a more substantial role in determining how athletes attenuate forces during landing.

Although the relationship was statistically significant, the weak correlation suggests that quadriceps strength explains only a small portion of the variability in landing mechanics, with other factors such as hip strength, trunk control, coordination, and task familiarity playing larger roles. These results differ modestly from existing evidence showing that in healthy populations thigh muscle strength and activation are poor predictors of knee flexion excursion during drop jump landing (2). Our findings are more consistent with prior studies using individuals post-ACL reconstruction showing greater quadriceps strength being associated with greater peak knee flexion angles during jump landings (3). Our results resemble the post ACL reconstruction cohorts more similarly even though all included participants were healthy. This may emphasize the effect of population differences between studies. Another consideration while interpreting our results is our outcome definition. Prior work with healthy athletes examined knee flexion excursion (2), while we examined peak knee flexion. Our data suggests that quadriceps strength may influence the maximum flexion achieved rather than the change from initial contact, which may depend on other factors beyond strength.

Stronger and more well-trained quadriceps may lead to greater peak knee flexion angles because the quadriceps eccentrically control knee flexion during landing. Based on our results, weaker quadriceps may be associated with reduced peak knee flexion during landing. This movement pattern reflects “stiffer” landing mechanics, characterized by extended knee positions, high quadriceps activity, and limited hamstring co-contraction. Stiffer landings could be a product of a more quadriceps-dominant landing strategy where relatively extended knee positions, high quadriceps activity, and limited hamstring co-contraction, could increase anterior tibial shear forces. This landing pattern may be associated with higher ground reaction forces and greater compressive loads at the tibiofemoral joint. The combination of the greater load with posterior tibial slope may further promote anterior tibial translation, causing an increased ACL injury risk. A more effective landing strategy may involve reduced stiffness with greater knee flexion. Our findings suggest stronger quadriceps could help generate the internal knee extensor moment required to control descent into deeper flexed positions, and therefore, potentially promote a softer landing, decreasing the risk of ACL injury. Also, it is important to note that there is a performance versus injury risk tradeoff. Stiffer landings allow athletes to move more quickly after they land because force is exerted over a shorter period, resulting in greater impulse and thus a greater change in momentum, which, given a constant mass, leads to higher velocity. It is also possible that greater strength can have diminishing returns in relation to injury risk. Specifically, there may be a threshold level of quadriceps strength required to achieve adequate knee flexion, beyond which additional strength gains may not meaningfully alter landing mechanics. Identifying this threshold is beneficial because it can guide strength benchmarks for rehabilitation or injury prevention screening.

A limitation of this study is the homogeneous sample of healthy adolescent athletes, which may limit generalizability to older or more highly trained populations. It is also possible that this landing task did not fully challenge athletes’ strength to assess if their strength dictated the way they landed. For instance, it is possible the task did not approach the athletes’ maximal or near-maximal capacity, limiting the extent to which strength would meaningfully influence movement strategy. Future studies could ameliorate this by including increased drop heights, adding external load, or incorporating more single-leg tasks to increase task demand. Another limitation is that quadriceps strength was assessed concentrically at a fixed, slow angular velocity, whereas landing requires rapid, high-velocity eccentric control, which may not be fully captured by this measure.

This study shows that there is a relationship between quadriceps strength and knee flexion angle from a vertical drop jump. These findings suggest that athletes with greater quadriceps strength may achieve greater knee flexion angles during landing, which has been associated with more favorable force attenuation strategies. Additionally, these findings support the continued inclusion of quadriceps strengthening as one component of ACL injury risk reduction programs, alongside targeted movement training to address landing mechanics.

## Data Availability

Data produced in the present study are available upon reasonable request to the authors.

